# The potential health impact of prospective Strep A vaccines: a modeling study

**DOI:** 10.1101/2022.08.01.22278298

**Authors:** Fiona Giannini, Jeffrey W. Cannon, Daniel Cadarette, David E. Bloom, Hannah C Moore, Jonathan Carapetis, Kaja Abbas

## Abstract

The World Health Organization published the preferred product characteristics for a Group A *Streptococcus* (Strep A) vaccine in 2018. Based on these parameters for the age of vaccination, vaccine efficacy, duration of protection from vaccine-derived immunity, and vaccination coverage, we developed a static cohort model to estimate the projected health impact of Strep A vaccination at the global, regional, and national levels and by country-income category. We used the model to analyse six strategic scenarios. Based on Strep A vaccine introduction between 2022 and 2034 for the primary scenario, we estimated vaccination at birth for 30 vaccinated cohorts could avert 2.5 billion episodes of pharyngitis, 345 million episodes of impetigo, 1.3 million episodes of invasive disease, 24 million episodes of cellulitis, and 6 million cases of rheumatic heart disease globally. Vaccination impact in terms of burden averted per fully vaccinated individual is highest in North America for cellulitis and in Sub-Saharan Africa for rheumatic heart disease.

## Introduction

Group A *Streptococcus* (Strep A) infection and direct sequelae are a major cause of morbidity and mortality at the global level, with more than half a million deaths annually^1–4^ attributable to Strep A. Strep A causes a broad spectrum of diseases, including pharyngitis, impetigo (skin infections), invasive disease, cellulitis, rheumatic heart disease (RHD), acute rheumatic fever (ARF), and acute post-streptococcal glomerulonephritis (APSGN / kidney disease). Children and young adults, particularly those in low- and middle-income countries, are the most affected by ARF and RHD,^5,6^ which impose substantial health burden in the form of morbidity and premature mortality due to cardiovascular impairment, as well as a substantial economic burden at the individual, household, and societal levels^7^. While relatively mild compared with RHD, cases and suspected cases of Strep A pharyngitis are major drivers of antibiotic use and misuse^8^, which can contribute to antimicrobial resistance in both Strep A and off-target (bystander) pathogens. While Strep A transmission occurs primarily through infections in the mucosae and skin, the specific dynamics of the transmission pathways are unknown.

The World Health Organization (WHO) published the preferred product characteristics for Strep A vaccines in 2018^9^, which describe the preferred parameters for vaccine indications, target population, data needs for safety and efficacy evaluation, research and development, and vaccination strategies. Strep A infection and disease spectrum is a WHO priority because of the unmet public health need for Strep A vaccines, assessment of technical feasibility, and suitability for use in low- and middle-income countries. While the WHO vision for Strep A vaccines and the near-term strategic goal is to demonstrate good safety and proof of efficacy of a candidate Strep A vaccine against pharyngitis and impetigo in children, the long-term strategic goal is to develop a safe, globally effective, and affordable Strep A vaccine to prevent acute Strep A infections (pharyngitis, impetigo, invasive disease, and cellulitis), avert associated antibiotic use and prevent secondary immune-mediated sequelae (APSGN, ARF, and RHD) and associated mortality.

Based on the WHO’s preferred product characteristics, we developed a mathematical model to assess the health impact of vaccination at the global, regional, and national levels and by country-income category. We used this Strep A vaccine impact model to analyse strategic scenarios for varied ages of vaccination (at birth or 5 years of age), vaccine efficacy, dynamics and duration of protection from vaccine-derived immunity, and vaccination coverage.

## Results

We used the vaccine efficacy assumptions (see Table 1) from the WHO preferred product characteristics for a Strep A vaccine and analysed six strategic scenarios (see Table 2) for varied years of vaccine introduction, coverage, and waning dynamics using the Strep A vaccine impact model (see Figure 1). Tables 3 and 4 and Figures 2–4 present the health impact of Strep A vaccination on lifetime disease burden averted among the vaccinated cohorts for the different scenarios.

**Table 1.** Vaccine efficacy. The vaccine efficacy assumptions are based on the WHO preferred product characteristics for the Strep A vaccine.

| Strep A disease state/sequelae | Vaccine efficacy (%) |
| --- | --- |
| Pharyngitis | 80 |
| Impetigo | 80 |
| Invasive disease | 70 |
| Cellulitis | 70 |
| Rheumatic heart disease | 50 |

**Table 2.**
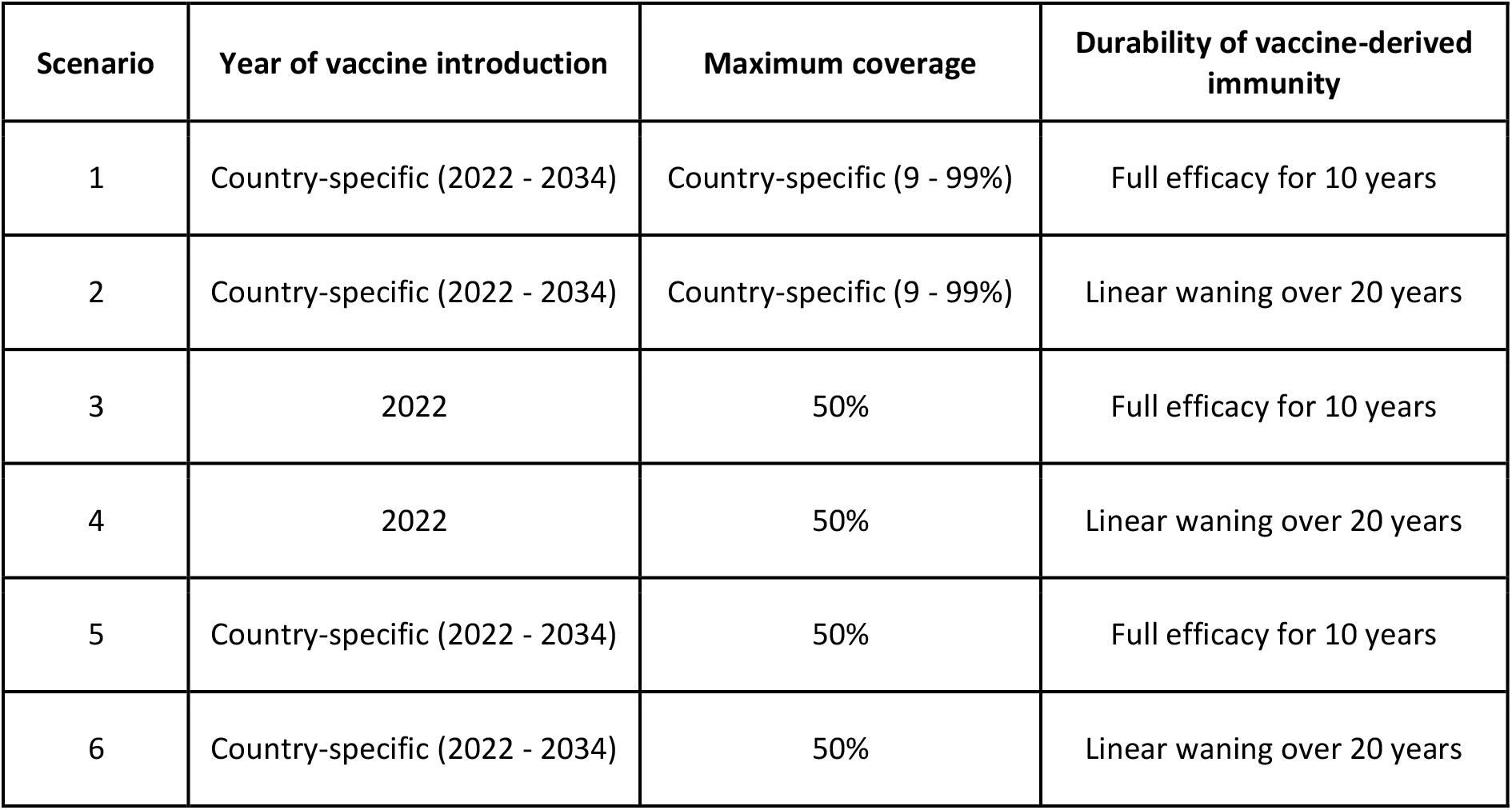
Vaccination scenarios. Potential vaccination scenarios for varying years of vaccine introduction, coverage, vaccine-derived immunity dynamics, and age of vaccination (at birth or 5 years of age).

**Table 3.** Vaccine impact at the regional and global levels. The vaccine impact on cases averted is presented at the regional (United Nations regions) and global levels for different scenarios, based on the lifetime health impact of vaccination at birth for 30 birth cohorts from year of vaccine introduction on Strep A disease burden (pharyngitis, impetigo, invasive disease, cellulitis, and rheumatic heart disease).

| UN regions | Scenarios | Fully vaccinated individuals (millions) | Cases averted through vaccination (thousands)<br>(range for scenarios 1-2 and 3-6) |  |  |  |  |
| --- | --- | --- | --- | --- | --- | --- | --- |
|  |  |  | Pharyngitis | Impetigo | Invasive disease | Cellulitis | Rheumatic heart disease |
| South Asia | 1-2* | 657 | (578,344, 606,913) | (76,156, 80,716) | (291, 310) | (3,880, 3,992) | (1,041, 1,466) |
|  | 3-6 | (381, 388) | (334,999, 357,905) | (44,117, 47,620) | (169, 183) | (2,246, 2,352) | (608, 870) |
| Europe & Central Asia | 1-2* | 226 | (199,636, 209,585) | (26,231, 27,796) | (100, 106) | (2,027, 2,104) | (102, 120) |
|  | 3-6 | (122, 124) | (107,661, 114,573) | (14,146, 15,197) | (54, 58) | (1,115, 1,171) | (53, 62) |
| Middle East & North Africa | 1-2* | 218 | (192,160, 201,701) | (25,266, 26,776) | (96, 102) | (1,253, 1,258) | (348, 407) |
|  | 3-6 | (121, 122) | (106,638, 112,917) | (14,025, 14,991) | (53, 57) | (693, 701) | (196, 232) |
| Sub-Saharan Africa | 1-2* | 918 | (799,501, 838,027) | (105,671, 112,075) | (407, 433) | (5,548, 5,635) | (3,635, 4,633) |
|  | 3-6 | (583, 607) | (505,846, 553,384) | (62,933, 74,061) | (258, 286) | (3,562, 3,777) | (2,285, 3,030) |
| Latin America & Caribbean | 1-2* | 184 | (162,482, 170,555) | (21,361, 22,637) | (81, 87) | (3,544, 3,949) | (413, 501) |
|  | 3-6 | (109, 113) | (95,898, 104,796) | (12,608, 13,913) | (48, 53) | (2,091, 2,435) | (244, 306) |
| East Asia & Pacific | 1-2* | 575 | (507,378, 532,590) | (66,695, 70,679) | (254, 270) | (3,261, 3,354) | (763, 860) |
|  | 3-6 | (317, 329) | (279,857, 304,513) | (36,790, 40,426) | (140, 155) | (1,837, 1,954) | (414, 483) |
| North America | 1-2* | 107 | (94,334, 99,039) | (12,395, 13,134) | (47, 50) | (4,145, 4,506) | (4, 4) |
|  | 3-6 | (58, 58) | (51,094, 53,879) | (6,714, 7,145) | (26, 27) | (2,245, 2,451) | (2, 2) |
| Global | 1-2* | 2,886 | (2,533,834, 2,658,410) | (333,775, 353,814) | (1,277, 1,359) | (23,657, 24,797) | (6,306, 7,991) |
|  | 3-6 | (1,690, 1,741) | (1,481,995, 1,601,967) | (195,332, 213,353) | (748, 820) | (13,789, 14,843) | (3,802, 4,985) |

\* Same number of fully vaccinated individuals for scenarios 1 and 2.

**Table 4.** Vaccine impact (deaths averted by vaccination at birth) at the regional and global levels. The vaccine impact on deaths averted (in thousands) is presented at the regional (United Nations regions) and global levels for different scenarios, based on the lifetime health impact of vaccination administered at birth for 30 birth cohorts from year of vaccine introduction on Strep A disease burden (invasive disease and rheumatic heart disease).

| UN regions | Scenarios | Fully vaccinated individuals (millions) | Deaths averted by vaccination at birth (thousands)<br>(range for scenarios 1-2 and 3-6) |  |
| --- | --- | --- | --- | --- |
|  |  |  | Invasive disease | Rheumatic heart disease |
| South Asia | 1-2* | 657 | (18, 19) | (313, 440) |
|  | 3-6 | (381, 388) | (10, 11) | (183, 261) |
| Europe & Central Asia | 1-2* | 226 | (6, 7) | (30, 36) |
|  | 3-6 | (122, 124) | (3, 4) | (16, 18) |
| Middle East & North Africa | 1-2* | 218 | (6, 6) | (103, 121) |
|  | 3-6 | (121, 122) | (3, 4) | (58, 69) |
| Sub-Saharan Africa | 1-2* | 918 | (25, 27) | (1,092, 1,392) |
|  | 3-6 | (583, 607) | (16, 18) | (686, 910) |
| Latin America & Caribbean | 1-2* | 184 | (5, 5) | (122, 148) |
|  | 3-6 | (109, 113) | (3, 3) | (72, 91) |
| East Asia & Pacific | 1-2* | 575 | (16, 17) | (229, 258) |
|  | 3-6 | (317, 329) | (9, 9) | (124, 145) |
| North America | 1-2* | 107 | (3, 3) | (0.11, 0.11) |
|  | 3-6 | (58, 58) | (2, 2) | (0.06, 0.06) |
| Global | 1-2* | 2,886 | (79, 83) | (1,889, 2,395) |
|  | 3-6 | (1,690, 1,741) | (46, 50) | (1,139, 1,494) |

\* Same number of fully vaccinated individuals for scenarios 1 and 2.

**Figure 1.**
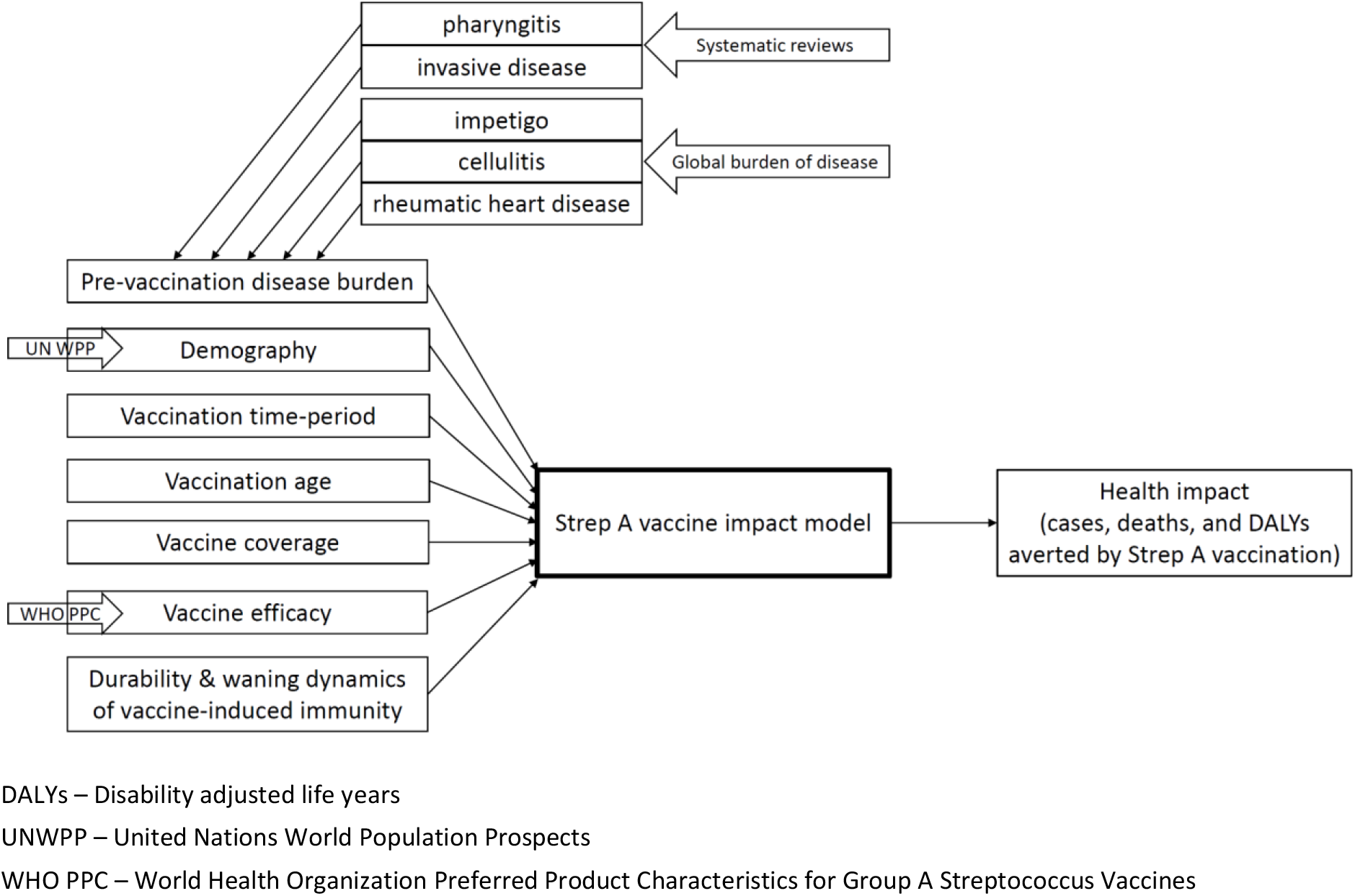
Strep A vaccine impact model. The static cohort model estimates the projected health impact of Strep A vaccination among the vaccinated cohorts over their lifetime in terms of burden averted for pharyngitis, invasive disease, impetigo, cellulitis, and rheumatic heart disease.

**Figure 2.**
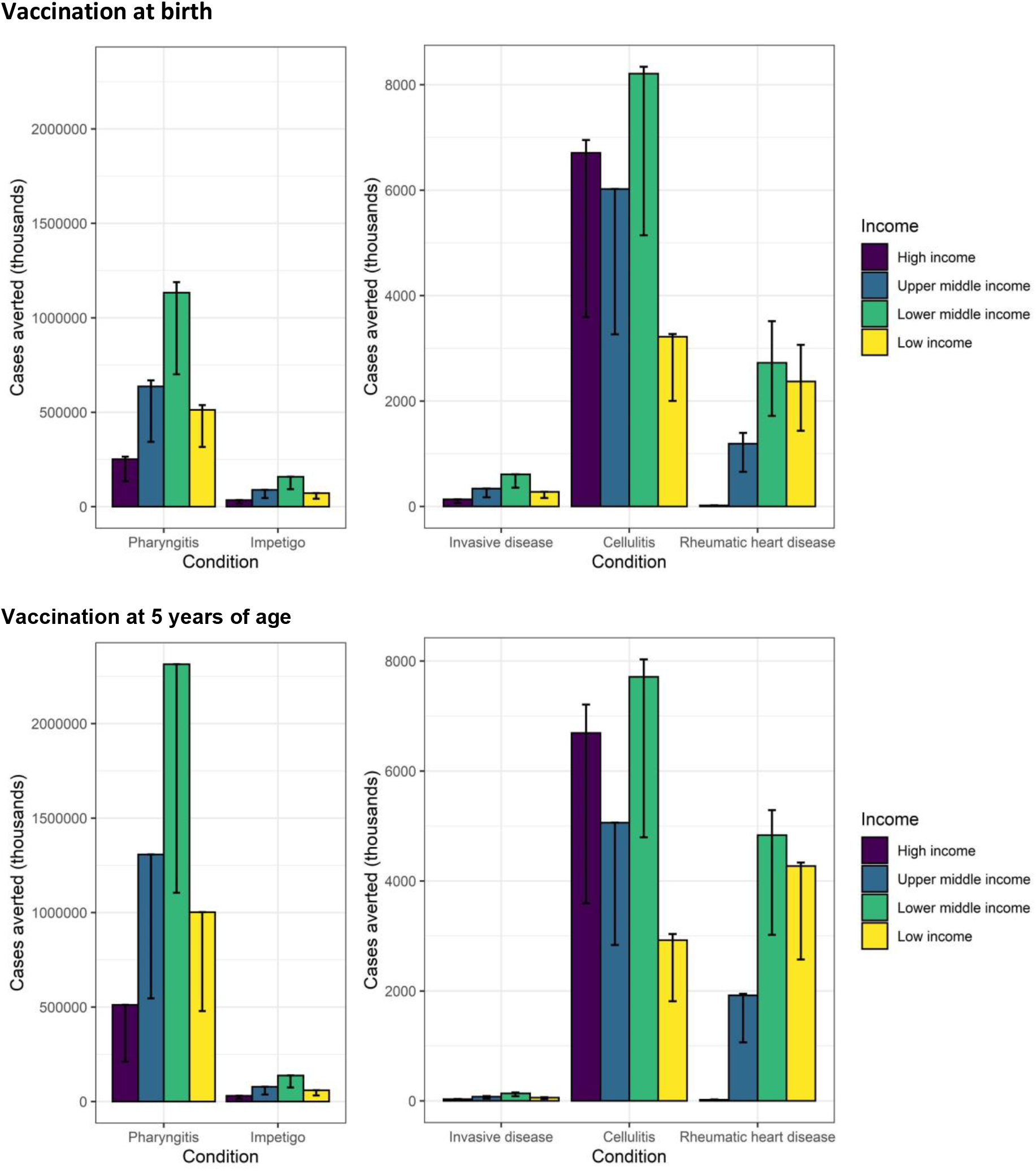
Vaccine impact at the country-income levels. The vaccine impact on cases averted (in thousands) is stratified by income levels of countries (World Bank income classification), based on the lifetime health impact of vaccination at birth or 5 years of age for 30 vaccinated cohorts from year of vaccine introduction on Strep A disease burden (pharyngitis, impetigo, invasive disease, cellulitis, and rheumatic heart disease). The vertical bars show the estimates for scenario 1, and the error bars show the range across scenarios 1-6. Note the differences in scale between the left panel (pharyngitis and impetigo) and the right panel (invasive, cellulitis and rheumatic heart disease).

### Vaccine impact on Strep A cases

Globally, we estimated that vaccination at birth for 30 cohorts assuming country-specific vaccine introduction between 2022 and 2034 under scenario 1 could avert 2.5 billion cases of pharyngitis, 354 million cases of impetigo, 1.4 million cases of invasive disease, 24 million cases of cellulitis, and 6 million cases of RHD during the vaccinated individuals’ lifetime (see Table 3 for scenario 1). This averages to 82 million cases of pharyngitis, 11.8 million cases of impetigo, 45,000 cases of invasive disease, 805,000 cases of cellulitis, and 210,000 cases of RHD averted per birth cohort.

Regionally, we estimated vaccination impact in terms of total cases averted to be highest in Sub-Saharan Africa (see Table 3) for pharyngitis, impetigo, invasive disease, cellulitis, and RHD. By income level, vaccination impact on Strep A cases averted was estimated to be highest in lower-middle-income countries (see Figure 2).

Vaccination impact in terms of cases averted per 1,000 vaccinated individuals was highest in North America for cellulitis and in Sub-Saharan Africa for RHD (see Figure 3). The vaccine impact metric of disease burden averted per 1,000 vaccinated individuals remains the same for any vaccination coverage in each scenario, with the caveat that the Strep A vaccine impact model includes only the direct effects of vaccination and excludes indirect herd effects. As only the direct effect of vaccination was included, the estimated health benefits of Strep A vaccination are likely to be conservative.

**Figure 3.**
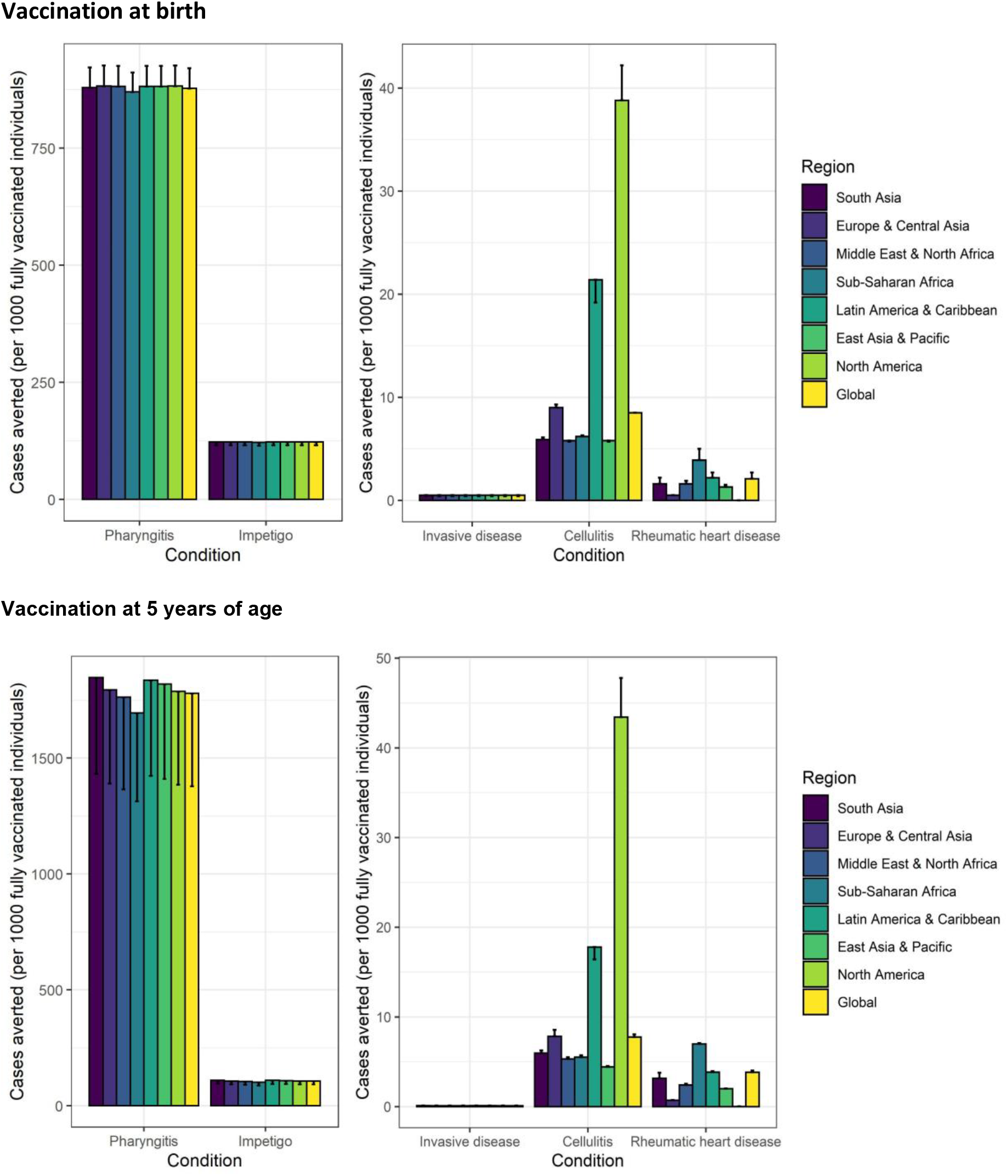
Vaccine impact at the regional and global levels. The vaccine impact on cases averted per 1,000 fully vaccinated individuals is stratified at the regional (United Nations regions) and global levels for different scenarios (estimate for scenario 1 and range across the six scenarios), based on the lifetime health impact of vaccination at birth or 5 years of age for 30 vaccinated cohorts from year of vaccine introduction on Strep A disease burden (pharyngitis, impetigo, invasive disease, cellulitis, and rheumatic heart disease). The vertical bars show the estimates for scenarios 1, 3, and 5 (which are equal), and the error bars show the estimates for scenarios 2, 4, and 6 (which are equal).

### Vaccine impact on Strep A-related deaths

Globally, we estimated that vaccination at birth for 30 cohorts under scenario 1 could avert 1.9 million deaths resulting from RHD and 79,000 deaths from invasive disease (see Table 4). Over 1 million of these global RHD deaths averted are in Sub-Saharan Africa (see Table 4).

### Vaccine impact on DALYs

Nationally (see Figure 4), we estimated the combined disability-adjusted life years (DALYs) per 1,000 vaccinated individuals averted due to a Strep A vaccine. This averted burden was highest in Tonga and Sub-Saharan African countries.

**Figure 4.**
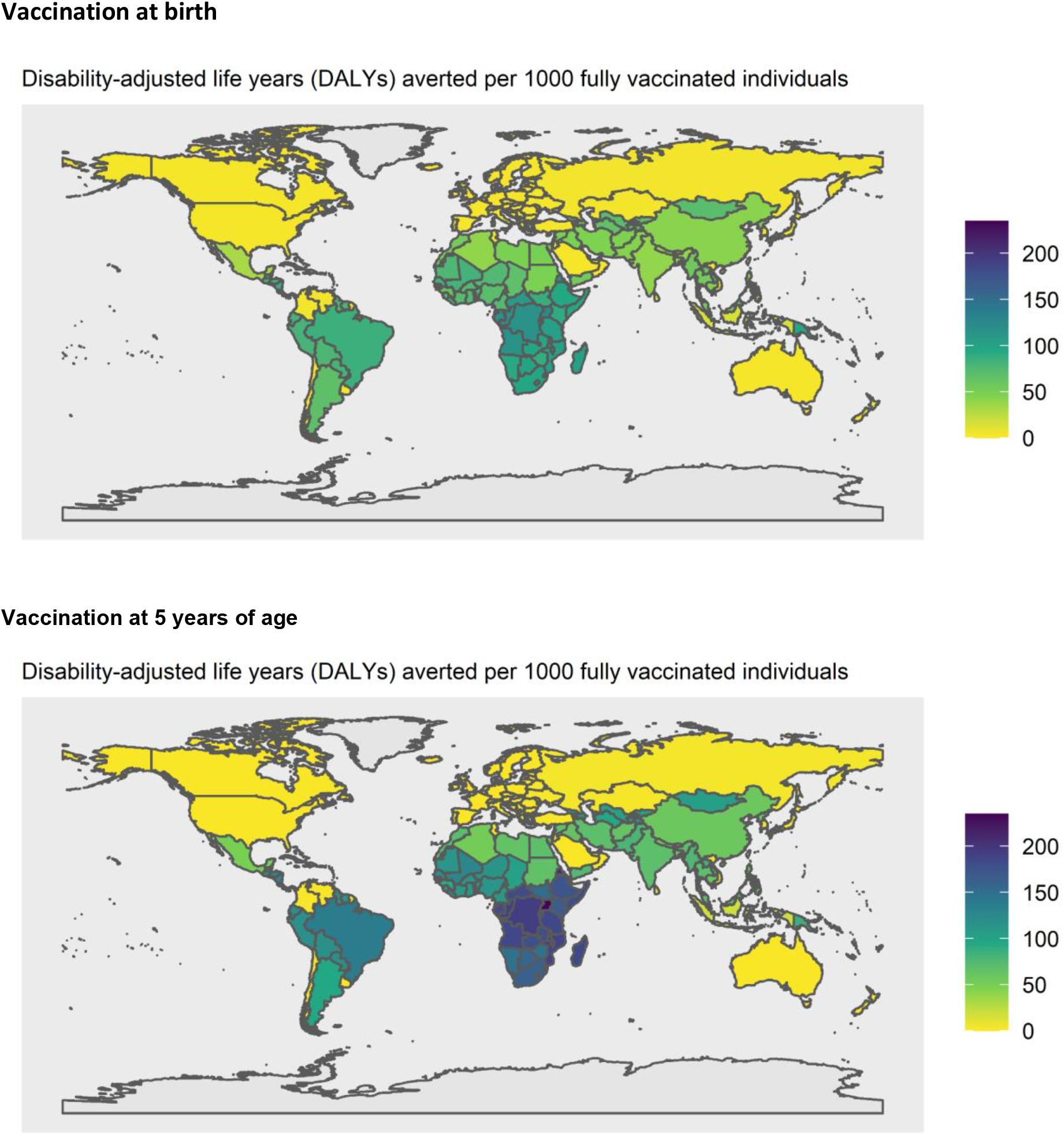
Vaccine impact at the national level. The vaccine impact on disability-adjusted life years (DALYs) averted per 1,000 fully vaccinated individuals is shown for 183 countries, based on the lifetime health impact of vaccination at birth or 5 years of age for 30 vaccinated cohorts from year of vaccine introduction on Strep A disease burden (pharyngitis, impetigo, invasive disease, cellulitis, and rheumatic heart disease) for scenario 1.

### Vaccine coverage levels and year of introduction

The numbers of fully vaccinated individuals were higher for scenarios 1-2 due to higher levels of vaccine coverage assumed for many of the countries (see Appendix) and hence a higher number of cases were averted in these scenarios compared with scenarios 3-6 (see Table 3). However, a delay in vaccine introduction occurs in scenarios 1–2 and 5–6 which leads to a corresponding delay in vaccination impact compared with scenarios 3–4 where Strep A vaccination was assumed to be introduced in all countries in 2022.

### Age of vaccination

Cases averted per 1,000 fully vaccinated individuals were higher for pharyngitis and RHD when vaccination was administered to children aged 5 years compared with vaccination at birth (see Figures 2-3). This is because the pre-vaccination burden (cumulative incidence) of pharyngitis and RHD is higher in the 10–20 years after 5 years of age than the cumulative incidence in the 10–20 years after birth among the 30 birth cohorts of 2022–2051.

Similarly, the combined DALYs averted resulting from a Strep A vaccine at the national level (see Figure 4) in countries in Sub-Saharan Africa, South America, and the Caribbean were higher when vaccination was administered at 5 years of age in comparison to vaccination at birth, primarily due to the higher number of RHD cases averted per 1,000 vaccinated individuals after 5 years of age.

## Discussion

We estimated the potential health impact of prospective Strep A vaccination at the global, regional, and national levels and by country-income category, using the Strep A vaccine impact model. Based on the WHO preferred product characteristics for Strep A vaccines,^9^ we developed this static cohort model to estimate the lifetime health benefits among the vaccinated cohorts, with a focus on Strep A disease burden averted for pharyngitis, impetigo, invasive disease, cellulitis, and RHD. We analysed strategic scenarios for vaccination at birth and 5 years of age and projected that around one-third of the annual global burden of RHD can be potentially averted by prospective Strep A vaccination. Regionally, vaccination impact in terms of burden averted per fully vaccinated individual is highest in North America for cellulitis and in Sub-Saharan Africa for RHD. By income level, total vaccine-avertable burdens for pharyngitis, impetigo, invasive disease, cellulitis, and RHD were highest in lower-middle-income countries, primarily due to the demographic effect of larger population sizes among the target ages for vaccination.

Our health impact estimates provide useful decision-making support for clinical development, interpretation of existing data, assessing needs for data collection to close critical evidence gaps, and policy decisions for prioritisation and implementation throughout the end-to-end continuum of discovery, development, and delivery of safe, effective, and affordable (prospective) Strep A vaccines. While the vaccine-avertable disease burden is an important public health priority, the high morbidity and mortality impact on the young adult population also translates into considerable productivity losses from an economic perspective^7,10^. This requires a broader global public health value proposition analysis to infer the end-to-end perspectives of vaccine development to uptake continuum^11–14^, and the global benefits of prospective Strep A vaccines are estimated to be highly positive^15^.

Our study has limitations. We estimated only the direct effect of Strep A vaccination and excluded the indirect effect on preventing pathogen transmission. Evidence on Strep A transmission dynamics would be valuable to develop a transmission dynamic model for estimating the total (direct and indirect) effects of vaccination. Such estimates would plausibly increase the impact of a Strep A vaccine, especially a vaccine administered in the first few years of life, given the peak incidences of severe invasive disease occurring in infants and older adults. Instead of endogenous modeling of transmission dynamics, the impact of indirect herd effects could be extrapolated by specifying a basic multiplier of the direct effect in the static cohort model, similar to the evaluation of *Haemophilus influenzae* type b, pneumococcal, and rotavirus vaccination^.16^

Evidence on the natural history of disease dynamics would be beneficial to simulate disease progression, such as the progression from acute rheumatic fever to RHD. We estimated vaccination impact on Strep A burden averted for pharyngitis, impetigo, invasive disease, cellulitis, and RHD but excluded the immune-mediated sequelae of acute rheumatic fever and acute post-streptococcal glomerulonephritis and the etiological pathway between Strep A infection and those diseases. While the exclusion of ARF and APSGN underestimates the impact of a vaccine that prevents those diseases, excluding the etiological pathway between infection and immune-mediated sequelae, including RHD, may overestimate the impact of vaccination at 5 years of age and underestimate the impact of infant vaccination. In our model, we assumed that vaccination in 5-year-old children would prevent incident cases of RHD occurring at age of 5 years. However, some proportion of those cases may be averted by an infant vaccination schedule that prevents the infections preceding ARF and RHD. Further epidemiology or immunology studies are required to model this scenario.

The vaccine impact model can be extended to include other Strep A disease states and sequelae to estimate the additional health benefits of vaccination in averting morbidity and mortality attributable to these conditions. The health benefits of vaccination in reducing Strep A infections could lower corresponding antibiotic use (to treat Strep A infections), and the model could capture this feature with the availability of quality data. The vaccine impact projections are based on a hypothetical vaccine that meets the WHO preferred product characteristics,^9^ and Strep A vaccines that attain licensure may have varied characteristics. The vaccine coverage assumptions are based on past coverage trends for *Haemophilus influenzae* type B vaccine (Hib3) or the third dose of diphtheria, tetanus, and pertussis vaccine, where Hib3 coverage values were unavailable, and coverage and scale-up of future Strep A vaccines may differ. Also, assumptions for the duration of protection and waning dynamics of vaccine-derived immunity may differ from future evidence generated from clinical trials and effectiveness studies.

In conclusion, based on the WHO preferred product characteristics for Strep A vaccines, we developed a Strep A vaccine impact model to estimate the health benefits of vaccination in terms of disease burden averted for pharyngitis, impetigo, invasive disease, cellulitis, and RHD at the global, regional, national, and country-income levels. The health impact estimates of Strep A vaccination are useful inputs for generating economic impact estimates based on cost-effectiveness analysis^17^ and for societal impact estimates based on benefit-cost analysis, thereby collating useful pieces of evidence towards the full value of vaccine assessment^11,18^ for Strep A vaccines.

## Methods

### Strep A vaccine impact model

We developed a static cohort model to estimate the lifetime health benefits of vaccination for 30 cohorts from the year of vaccine introduction (2022-2034) on Strep A disease burden (pharyngitis, impetigo, invasive disease, cellulitis, and RHD) in terms of episodes/cases, deaths, and disability-adjusted life years (DALYs) averted by vaccination (see Figure 1). The reduction in disease burden is in direct proportion to vaccine efficacy, vaccine coverage, and vaccine-derived immunity.

The pre-vaccination disease burden is based on country- and age-specific incidence rates for cellulitis and RHD and global age-specific prevalence for impetigo from the 2019 Global Burden of Disease (GBD) study^19^. For pharyngitis and invasive disease, the pre-vaccination disease burden is based on global age-specific rates from systematic reviews conducted as part of the Strep A Vaccine Global Consortium (SAVAC) project. Mortality risk was limited to 28 days from hospitalisation for invasive disease and to 10 years from disease onset for RHD. Country- and age-specific rates of Strep A burden were assumed to remain constant in the future. We did not include APSGN and ARF in our analysis due to data limitations on prevalent burden estimates.

Demographic estimates for the country, year and age-specific population, all-cause mortality rates, and remaining life expectancy are based on the 2019 United Nations World Population Prospects^20^. The model uses non-sex specific projected (2020 - 2100) interpolated age-specific (0 - 99 years) population and age-grouped (covering the same age range) all-cause mortality probabilities and remaining life expectancy estimates. Age groups for all-cause mortality and life expectancy are in 5-year bands, apart from 0 – 4 years (age 0 is in a separate group), assuming uniformity within groups for mortality and remaining life expectancy. Any projection of lifetime burden that went beyond 2100 assumed the same population, all-cause mortality, and remaining life expectancy values as for 2100. The country- and age-specific population numbers were used to estimate the population at age of vaccination, and then all-cause mortality probabilities were used to estimate the modelled population at each age over a cohort’s lifetime.

Disability weights used for the calculation of years lived with disability are from the GBD study,^21^ and YLD (years lived with disability) was attributed to the years of prevalence. The duration for pharyngitis, impetigo, invasive disease, and cellulitis was estimated to be 5 days, 15.5 days, 10 days, and 16.4 days respectively, based on the GBD reported prevalence divided by incidence.^19^ The duration for RHD was assumed to be the remaining life expectancy from the onset of the condition.

The vaccine efficacy assumptions are based on the WHO preferred product characteristics for Strep A vaccine, as shown in Table 1.^9^ The waning dynamics of vaccine-derived immunity were modelled in two ways: (i) vaccine-induced immune protection at maximum efficacy for 10 years and null thereafter and (ii) waning linearly with an annual reduction in efficacy equivalent to 5% of maximum efficacy for 20 years and null thereafter (i.e., waning to 50% of maximum efficacy after 10 years). The year of vaccine introduction was assumed to be 2022 or country-specific ranging from 2022 to 2034, with initial coverage at 10% of maximum coverage. The vaccine coverage was assumed to scale up linearly during the first 10 years after introduction to reach either a maximum of 50% coverage for all countries or a country-specific coverage ranging from 9% to 99%. Country-specific coverage values and year of introduction (see Appendix) are based on past trends for Hib3 or the third dose of diphtheria, tetanus, and pertussis vaccine where Hib3 values were unavailable.

Using the model, we analysed six potential scenarios for varied years of vaccine introduction, coverage, and waning dynamics (see Table 2), with vaccination at birth or 5 years of age. We modelled these scenarios for 183 countries (see Table A5) and aggregated results at the global, regional, and country-income levels.

While these vaccination scenarios are hypothetical, the pragmatic vaccination scenarios will become clear during the vaccine introduction discussions at the national (including national immunization technical advisory groups), regional (including regional immunization technical advisory groups), and global (including WHO Strategic Advisory Group of Experts on immunization) levels. However, the vaccine impact metric of disease burden averted per fully vaccinated individual remains the same for any vaccination coverage for a given scenario, with the caveat that the Strep A vaccine impact model includes only the direct effects of vaccination and excludes indirect herd effects.

### R package and Shiny web application

We developed the Strep A vaccine impact model using the R statistical software^22^ and included a user-friendly R Shiny web application. The program code and data for the vaccine impact model is available as an R package, *GASImpactModel* (https://github.com/fionagi/GASImpactModel) and modeling analysis can be conducted through the R Shiny web application (https://github.com/fionagi/GASImpactModel_App). Through the web application, the impact of a selected vaccination scenario can be visualized for any of 205 countries. The app shows the predicted lifetime health benefits from age of vaccination associated with the vaccination of multiple cohorts during the period selected. The vaccine impact model can estimate the health benefits of vaccination by calendar year, birth year, and year of vaccination.^23^

## Data Availability

All data produced in the present study are available upon reasonable request to the authors

## Contribution

FG, JWC, and KA conceptualized and designed the study. FG and JWC compiled the data sets and conducted the analysis. FG developed the Strep A vaccine impact model and software. DC, DEB, HCM, and JC advised on modeling framework and public health implications. FG and KA wrote the first draft. All authors contributed with critical input, reviewing and editing of the manuscript, and have approved the final version.

## Acknowledgements

We thank Kate Miller (Telethon Kids Institute) for helpful discussions.

## Appendix

**Table A1.**
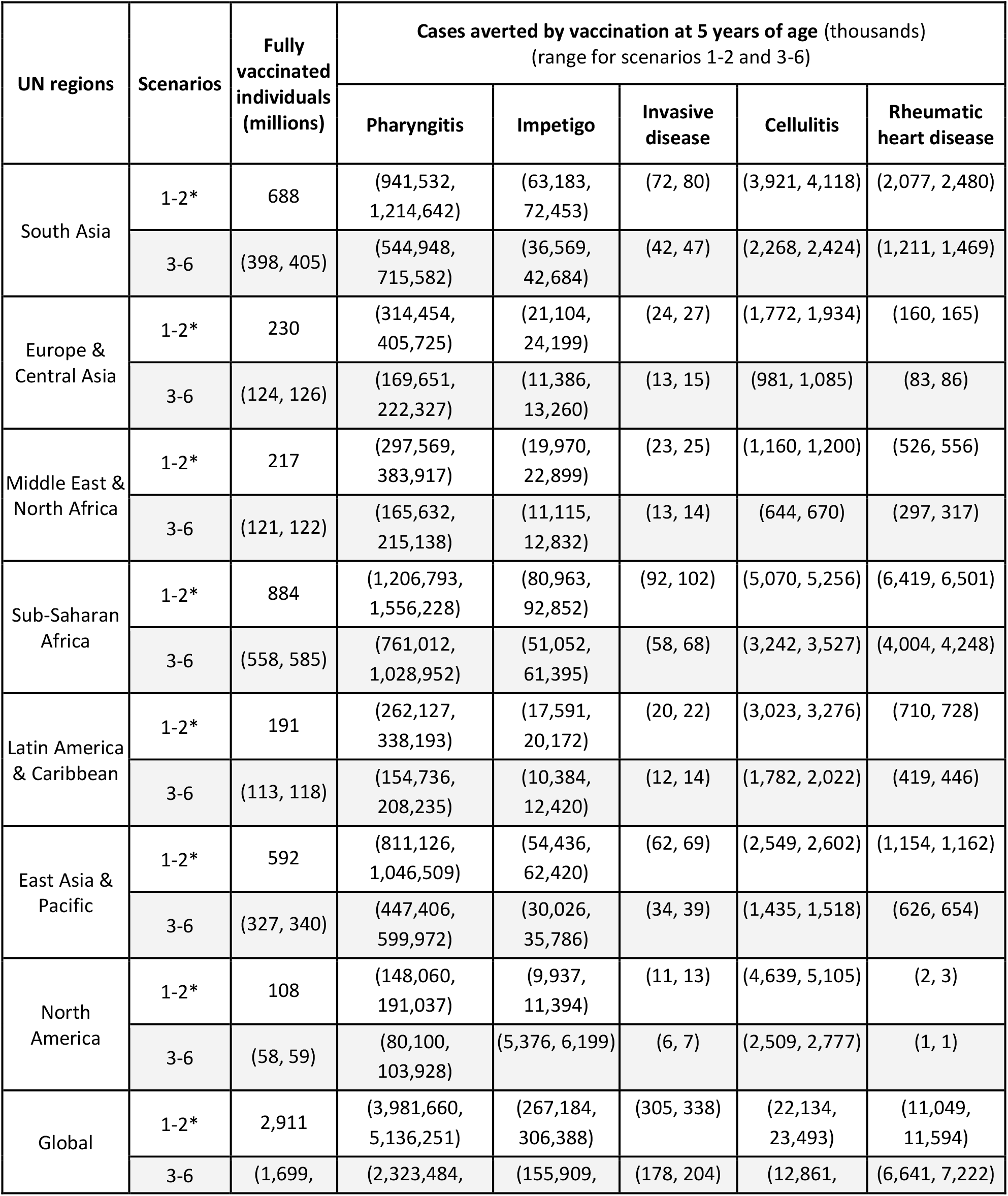
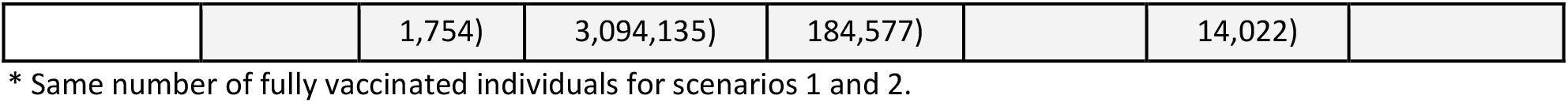
Vaccine impact (cases averted by vaccination at 5 years of age) at the regional and global levels. The vaccine impact on cases averted is presented at the regional (United Nations regions) and global levels for different scenarios, based on the lifetime health impact of vaccination administered at 5 years of age for 30 vaccinated cohorts on Strep A disease burden (pharyngitis, impetigo, invasive disease, cellulitis, and rheumatic heart disease).

**Table A2.** Vaccine impact (DALYs averted by vaccination at birth) at the regional and global levels. The vaccine impact on DALYs averted (in thousands) is presented at the regional (United Nations regions) and global levels for different scenarios, based on the lifetime health impact of vaccination administered at birth for 30 vaccinated cohorts on Strep A disease burden (pharyngitis, impetigo, invasive disease, cellulitis, and rheumatic heart disease).

| UN regions | Scenarios | Fully vaccinated individuals (millions) | DALYs averted by vaccination at birth (thousands)<br>(range for scenarios 1-2 and 3-6) |  |  |  |  |
| --- | --- | --- | --- | --- | --- | --- | --- |
|  |  |  | Pharyngitis | Impetigo | Invasive disease | Cellulitis | Rheumatic heart disease |
| South Asia | 1-2* | 657 | (206, 216) | (19, 21) | (1,321, 1,400) | (9, 9) | (24,398, 32,854) |
|  | 3-6 | (381, 388) | (119, 127) | (11, 12) | (764, 820) | (5, 5) | (14,229, 19,354) |
| Europe & Central Asia | 1-2* | 226 | (71, 75) | (7, 7) | (495, 525) | (5, 5) | (2,387, 2,720) |
|  | 3-6 | (122, 124) | (38, 41) | (4, 4) | (267, 284) | (3, 3) | (1,233, 1,413) |
| Middle East & North Africa | 1-2* | 218 | (68, 72) | (6, 7) | (458, 485) | (3, 3) | (8,394, 9,507) |
|  | 3-6 | (121, 122) | (38, 40) | (4, 4) | (251, 271) | (2, 2) | (4,678, 5,410) |
| Sub-Saharan Africa | 1-2* | 918 | (285, 298) | (27, 29) | (1,710, 1,813) | (13, 13) | (79,908, 98,237) |
|  | 3-6 | (583, 607) | (180, 197) | (17, 19) | (1,068, 1,191) | (8, 9) | (49,637, 63,933) |
| Latin America & Caribbean | 1-2* | 184 | (58, 61) | (5, 6) | (398, 422) | (8, 9) | (10,341, 12,141) |
|  | 3-6 | (109, 113) | (34, 37) | (3, 4) | (235, 256) | (5, 6) | (6,095, 7,338) |
| East Asia & Pacific | 1-2* | 575 | (181, 190) | (17, 18) | (1,240, 1,314) | (7, 8) | (19,143, 21,000) |
|  | 3-6 | (317, 329) | (100, 108) | (9, 10) | (682, 739) | (4, 4) | (10,356, 11,603) |
| North America | 1-2* | 107 | (34, 35) | (3, 3) | (239, 253) | (9, 10) | (22, 22) |
|  | 3-6 | (58, 58) | (18, 19) | (2, 2) | (128, 138) | (5, 6) | (12, 12) |
| Global | 1-2* | 2,886 | (902, 946) | (85, 90) | (5,862, 6,212) | (54, 57) | (144,593, 176,481) |
|  | 3-6 | (1,690, 1,741) | (527, 570) | (50, 54) | (3,396, 3,699) | (32, 34) | (86,241, 109,064) |

\* Same number of fully vaccinated individuals for scenarios 1 and 2.

**Table A3.**
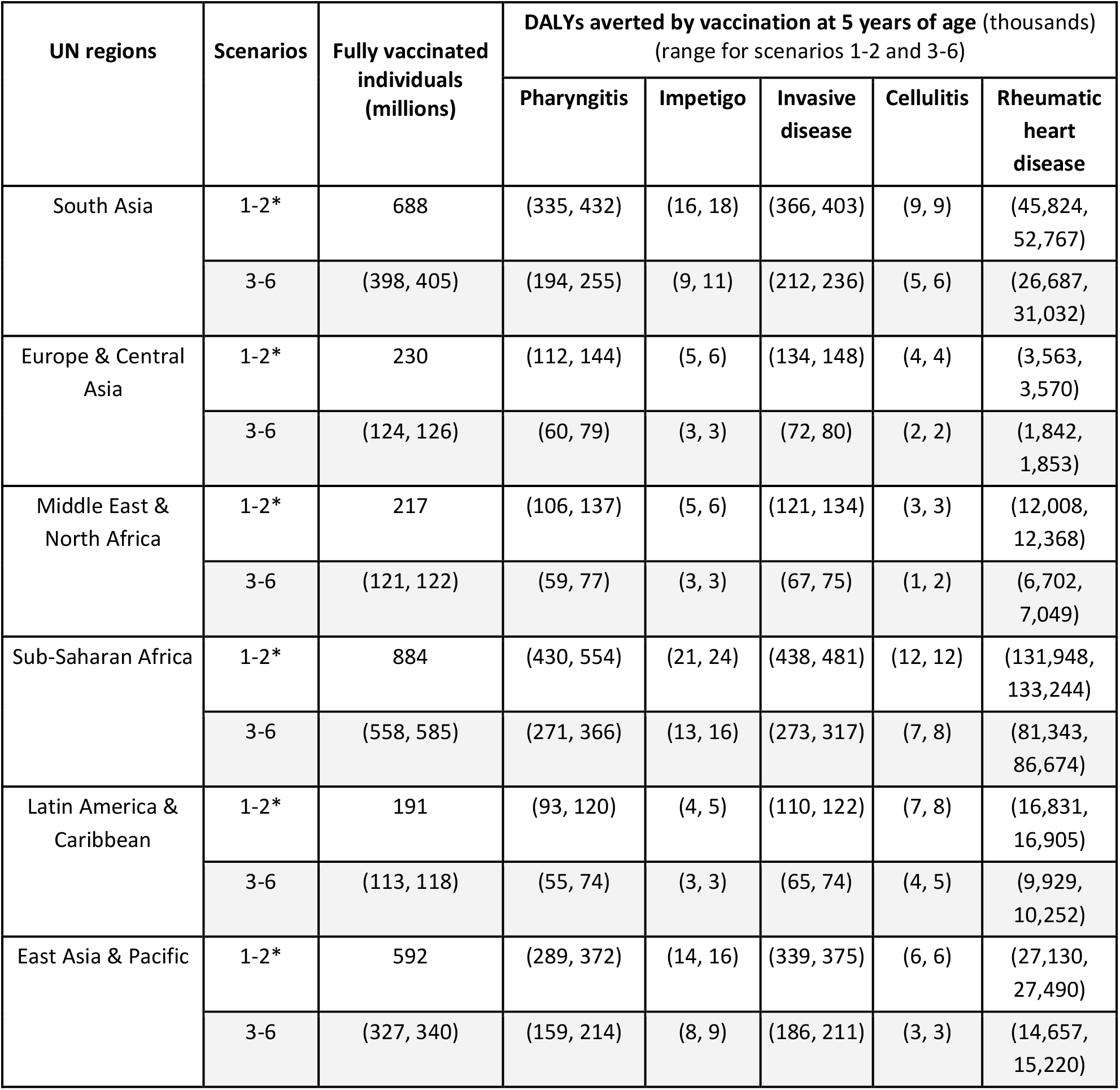
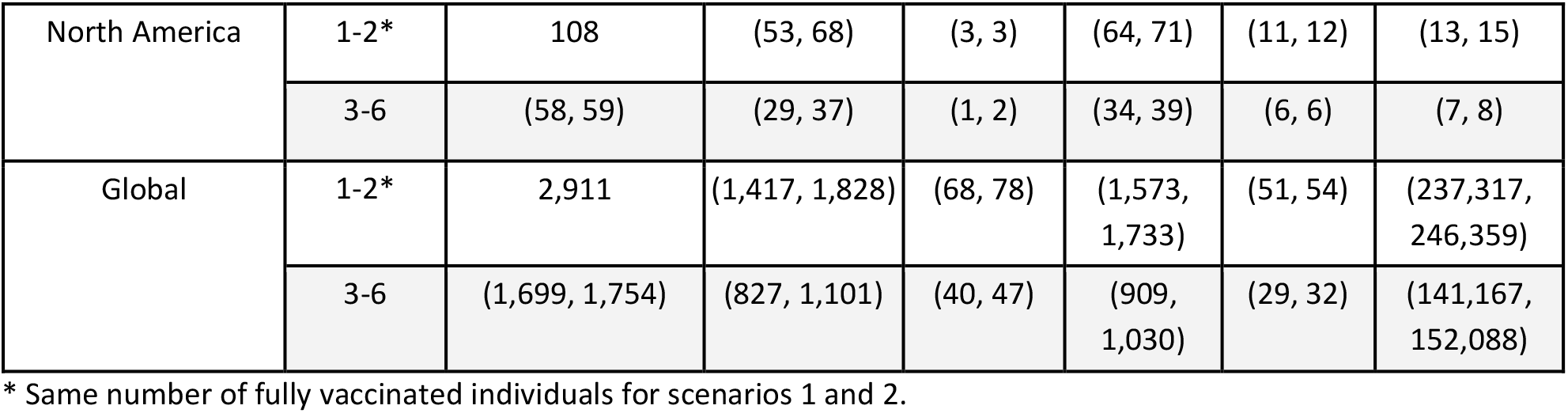
Vaccine impact (DALYs averted by vaccination at 5 years of age) at the regional and global levels. The vaccine impact on DALYs averted (in thousands) is presented at the regional (United Nations regions) and global levels for different scenarios, based on the lifetime health impact of vaccination administered at 5 years of age for 30 vaccinated cohorts on Strep A disease burden (pharyngitis, impetigo, invasive disease, cellulitis, and rheumatic heart disease).

| UN regions | Scenarios | Fully vaccinated individuals (millions) | DALYs averted by vaccination at 5 years of age (thousands)<br>(range for scenarios 1-2 and 3-6) |  |  |  |  |
| --- | --- | --- | --- | --- | --- | --- | --- |
|  |  |  | Pharyngitis | Impetigo | Invasive disease | Cellulitis | Rheumatic heart disease |
| South Asia | 1-2* | 688 | (335, 432) | (16, 18) | (366, 403) | (9, 9) | (45,824, 52,767) |
|  | 3-6 | (398, 405) | (194, 255) | (9, 11) | (212, 236) | (5, 6) | (26,687, 31,032) |
| Europe & Central Asia | 1-2* | 230 | (112, 144) | (5, 6) | (134, 148) | (4, 4) | (3,563, 3,570) |
|  | 3-6 | (124, 126) | (60, 79) | (3, 3) | (72, 80) | (2, 2) | (1,842, 1,853) |
| Middle East & North Africa | 1-2* | 217 | (106, 137) | (5, 6) | (121, 134) | (3, 3) | (12,008, 12,368) |
|  | 3-6 | (121, 122) | (59, 77) | (3, 3) | (67, 75) | (1, 2) | (6,702, 7,049) |
| Sub-Saharan Africa | 1-2* | 884 | (430, 554) | (21, 24) | (438, 481) | (12, 12) | (131,948, 133,244) |
|  | 3-6 | (558, 585) | (271, 366) | (13, 16) | (273, 317) | (7, 8) | (81,343, 86,674) |
| Latin America & Caribbean | 1-2* | 191 | (93, 120) | (4, 5) | (110, 122) | (7, 8) | (16,831, 16,905) |
|  | 3-6 | (113, 118) | (55, 74) | (3, 3) | (65, 74) | (4, 5) | (9,929, 10,252) |
| East Asia & Pacific | 1-2* | 592 | (289, 372) | (14, 16) | (339, 375) | (6, 6) | (27,130, 27,490) |
|  | 3-6 | (327, 340) | (159, 214) | (8, 9) | (186, 211) | (3, 3) | (14,657, 15,220) |
| North America | 1-2* | 108 | (53, 68) | (3, 3) | (64, 71) | (11, 12) | (13, 15) |
|  | 3-6 | (58, 59) | (29, 37) | (1, 2) | (34, 39) | (6, 6) | (7, 8) |
| Global | 1-2* | 2,911 | (1,417, 1,828) | (68, 78) | (1,573, 1,733) | (51, 54) | (237,317, 246,359) |
|  | 3-6 | (1,699, 1,754) | (827, 1,101) | (40, 47) | (909, 1,030) | (29, 32) | (141,167, 152,088) |

\* Same number of fully vaccinated individuals for scenarios 1 and 2.

**Table A4.** Vaccine impact (deaths averted by vaccination at 5 years of age) at the regional and global levels. The vaccine impact on deaths averted (in thousands) is presented at the regional (United Nations regions) and global levels for different scenarios, based on the lifetime health impact of vaccination administered at 5 years of age for 30 vaccinated cohorts on Strep A disease burden (invasive disease and rheumatic heart disease).

| UN regions | Scenarios | Fully vaccinated individuals (millions) | Deaths averted by vaccination at 5 years of age (thousands)<br>(range for scenarios 1-2 and 3-6) |  |
| --- | --- | --- | --- | --- |
|  |  |  | Invasive disease | Rheumatic heart disease |
| South Asia | 1-2* | 688 | (5, 6) | (624, 745) |
|  | 3-6 | (398, 405) | (3, 4) | (364, 441) |
| Europe & Central Asia | 1-2* | 230 | (2, 2) | (48, 49) |
|  | 3-6 | (124, 126) | (1, 1) | (25, 25) |
| Middle East & North Africa | 1-2* | 217 | (2, 2) | (156, 165) |
|  | 3-6 | (121, 122) | (1, 1) | (88, 94) |
| Sub-Saharan Africa | 1-2* | 884 | (7, 8) | (1,928, 1,953) |
|  | 3-6 | (558, 585) | (4, 5) | (1,203, 1,276) |
| Latin America & Caribbean | 1-2* | 191 | (1, 2) | (210, 216) |
|  | 3-6 | (113, 118) | (1, 1) | (124, 132) |
| East Asia & Pacific | 1-2* | 592 | (5, 5) | (346, 349) |
|  | 3-6 | (327, 340) | (3, 3) | (188, 196) |
| North America | 1-2* | 108 | (1, 1) | (0.07, 0.08) |
|  | 3-6 | (58, 59) | (0.45, 0.52) | (0.04, 0.04) |
| Global | 1-2* | 2,911 | (23, 26) | (3,313, 3,476) |
|  | 3-6 | (1,699, 1,754) | (13, 15) | (1,992, 2,166) |

\* Same number of fully vaccinated individuals for scenarios 1 and 2.

**Table A5.** Country-specific coverage and year of introduction for Strep A vaccination.

| **Country** | **Vaccine coverage (%)** | **Vaccine introduction (year)** |
| --- | --- | --- |
| Afghanistan | 66 | 2030 |
| Albania | 99 | 2025 |
| Algeria | 91 | 2026 |
| Angola | 59 | 2027 |
| Antigua and Barbuda | 95 | 2027 |
| Argentina | 86 | 2027 |
| Armenia | 92 | 2022 |
| Australia | 94 | 2025 |
| Austria | 85 | 2030 |
| Azerbaijan | 95 | 2026 |
| Bahamas, The | 90 | 2027 |
| Bahrain | 99 | 2027 |
| Bangladesh | 98 | 2025 |
| Barbados | 95 | 2026 |
| Belarus | 9 | 2030 |
| Belgium | 97 | 2027 |
| Belize | 96 | 2028 |
| Benin | 76 | 2026 |
| Bhutan | 97 | 2028 |
| Bolivia | 83 | 2027 |
| Bosnia and Herzegovina | 62 | 2032 |
| Botswana | 95 | 2022 |
| Brazil | 83 | 2027 |
| Brunei Darussalam | 99 | 2030 |
| Bulgaria | 92 | 2027 |
| Burkina Faso | 91 | 2022 |
| Burundi | 90 | 2022 |
| Cabo Verde | 99 | 2025 |
| Cambodia | 92 | 2026 |
| Cameroon | 79 | 2027 |
| Canada | 91 | 2025 |
| Central African Republic | 47 | 2028 |
| Chad | 41 | 2030 |
| Chile | 95 | 2029 |
| China | 99 | 2028 |
| Colombia | 92 | 2027 |
| Comoros | 91 | 2025 |
| Congo, Dem. Rep. | 81 | 2028 |
| Congo, Rep. | 75 | 2025 |
| Costa Rica | 94 | 2025 |
| Côte d'Ivoire | 82 | 2025 |
| Croatia | 94 | 2030 |
| Cuba | 99 | 2028 |
| Cyprus | 97 | 2029 |
| Czech Republic / Czechia | 94 | 2030 |
| Denmark | 97 | 2029 |
| Djibouti | 84 | 2025 |
| Dominican Republic | 90 | 2027 |
| Ecuador | 85 | 2027 |
| Egypt, Arab Rep. | 95 | 2028 |
| El Salvador | 81 | 2027 |
| Equatorial Guinea | 25 | 2030 |
| Eritrea | 95 | 2022 |
| Estonia | 92 | 2027 |
| Eswatini | 90 | 2022 |
| Ethiopia | 72 | 2027 |
| Fiji | 99 | 2022 |
| Finland | 91 | 2027 |
| France | 95 | 2029 |
| Gabon | 70 | 2028 |
| Gambia, The | 93 | 2025 |
| Georgia | 93 | 2025 |
| Germany | 92 | 2027 |
| Ghana | 97 | 2022 |
| Greece | 99 | 2027 |
| Grenada | 96 | 2028 |
| Guatemala | 86 | 2027 |
| Guinea | 45 | 2030 |
| Guinea-Bissau | 88 | 2025 |
| Guyana | 95 | 2025 |
| Haiti | 64 | 2028 |
| Honduras | 90 | 2027 |
| Hungary | 99 | 2029 |
| Iceland | 91 | 2029 |
| India | 89 | 2025 |
| Indonesia | 79 | 2032 |
| Iran, Islamic Rep. | 99 | 2027 |
| Iraq | 84 | 2027 |
| Ireland | 94 | 2027 |
| Israel | 98 | 2027 |
| Italy | 94 | 2027 |
| Jamaica | 98 | 2027 |
| Japan | 99 | 2029 |
| Jordan | 96 | 2031 |
| Kazakhstan | 98 | 2029 |
| Kenya | 92 | 2022 |
| Kiribati | 95 | 2022 |
| Korea, Dem. People's Rep. | 97 | 2028 |
| Korea, Rep. | 98 | 2028 |
| Kuwait | 99 | 2027 |
| Kyrgyz Republic | 92 | 2025 |
| Lao PDR | 68 | 2028 |
| Latvia | 96 | 2027 |
| Lebanon | 85 | 2031 |
| Lesotho | 93 | 2025 |
| Liberia | 84 | 2025 |
| Libya | 97 | 2027 |
| Lithuania | 92 | 2027 |
| Luxembourg | 99 | 2027 |
| Madagascar | 75 | 2025 |
| Malawi | 92 | 2022 |
| Malaysia | 99 | 2028 |
| Maldives | 99 | 2028 |
| Mali | 71 | 2025 |
| Malta | 97 | 2030 |
| Mauritania | 81 | 2025 |
| Mauritius | 97 | 2025 |
| Mexico | 88 | 2030 |
| Micronesia, Fed. Sts. | 59 | 2025 |
| Moldova | 92 | 2027 |
| Mongolia | 99 | 2026 |
| Montenegro | 87 | 2033 |
| Morocco | 99 | 2025 |
| Mozambique | 80 | 2025 |
| Myanmar | 91 | 2026 |
| Namibia | 89 | 2025 |
| Nepal | 91 | 2026 |
| Netherlands | 93 | 2029 |
| New Zealand | 92 | 2025 |
| Nicaragua | 98 | 2025 |
| Niger | 89 | 2025 |
| Nigeria | 57 | 2028 |
| North Macedonia | 91 | 2030 |
| Norway | 96 | 2027 |
| Oman | 99 | 2029 |
| Pakistan | 75 | 2027 |
| Panama | 88 | 2027 |
| Papua New Guinea | 61 | 2027 |
| Paraguay | 88 | 2027 |
| Peru | 84 | 2027 |
| Philippines | 65 | 2032 |
| Poland | 95 | 2029 |
| Portugal | 99 | 2029 |
| Qatar | 98 | 2027 |
| Romania | 86 | 2032 |
| Russian Federation | 97 | 2030 |
| Rwanda | 97 | 2022 |
| Samoa | 34 | 2030 |
| São Tomé and Príncipe | 95 | 2022 |
| Saudi Arabia | 96 | 2027 |
| Senegal | 82 | 2022 |
| Serbia | 96 | 2029 |
| Seychelles | 99 | 2025 |
| Sierra Leone | 90 | 2022 |
| Singapore | 96 | 2029 |
| Slovak Republic / Slovakia | 96 | 2029 |
| Slovenia | 93 | 2029 |
| Solomon Islands | 85 | 2024 |
| Somalia | 42 | 2030 |
| South Africa | 74 | 2024 |
| South Sudan | 49 | 2030 |
| Spain | 94 | 2029 |
| Sri Lanka | 99 | 2030 |
| St. Lucia | 95 | 2027 |
| St. Vincent and the Grenadines | 97 | 2028 |
| Sudan | 93 | 2025 |
| Suriname | 95 | 2030 |
| Sweden | 97 | 2029 |
| Switzerland | 95 | 2029 |
| Syrian Arab Republic | 48 | 2033 |
| Tajikistan | 96 | 2026 |
| Tanzania | 98 | 2025 |
| Thailand | 97 | 2030 |
| Timor-Leste | 83 | 2030 |
| Togo | 88 | 2025 |
| Tonga | 81 | 2024 |
| Trinidad and Tobago | 99 | 2026 |
| Tunisia | 97 | 2030 |
| Turkey | 98 | 2029 |
| Turkmenistan | 99 | 2026 |
| Uganda | 93 | 2022 |
| Ukraine | 39 | 2033 |
| United Arab Emirates | 99 | 2025 |
| United Kingdom | 94 | 2026 |
| United States | 92 | 2027 |
| Uruguay | 91 | 2029 |
| Uzbekistan | 98 | 2025 |
| Vanuatu | 85 | 2025 |
| Venezuela, RB | 60 | 2034 |
| Vietnam | 75 | 2033 |
| Yemen, Rep. | 65 | 2027 |
| Zambia | 90 | 2025 |
| Zimbabwe | 89 | 2022 |

**Figure A1.**
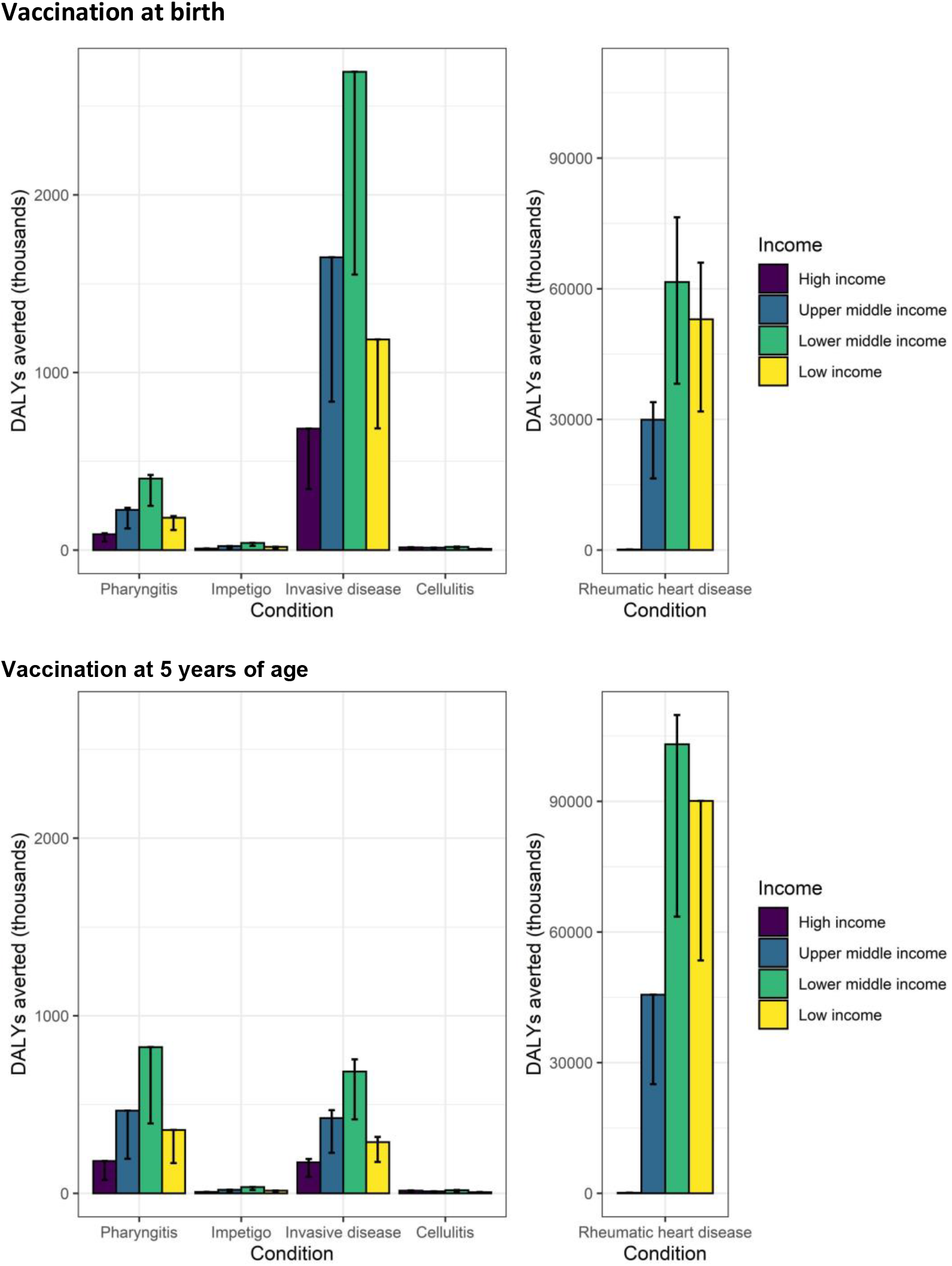
Vaccine impact (DALYs averted) at the country-income level. The vaccine impact on DALYs averted (in thousands) is stratified by income levels of countries (World Bank income classification), based on the lifetime health impact of vaccination at birth or 5 years of age for 30 vaccinated cohorts on Strep A disease burden (pharyngitis, impetigo, invasive disease, cellulitis, and rheumatic heart disease). The vertical bars show the estimates for scenario 1, and the error bars show the range across scenarios 1-6. Note the differences in scale between the left panel (pharyngitis, impetigo and cellulitis) and the right panel (invasive and rheumatic heart disease).

**Figure A2.**
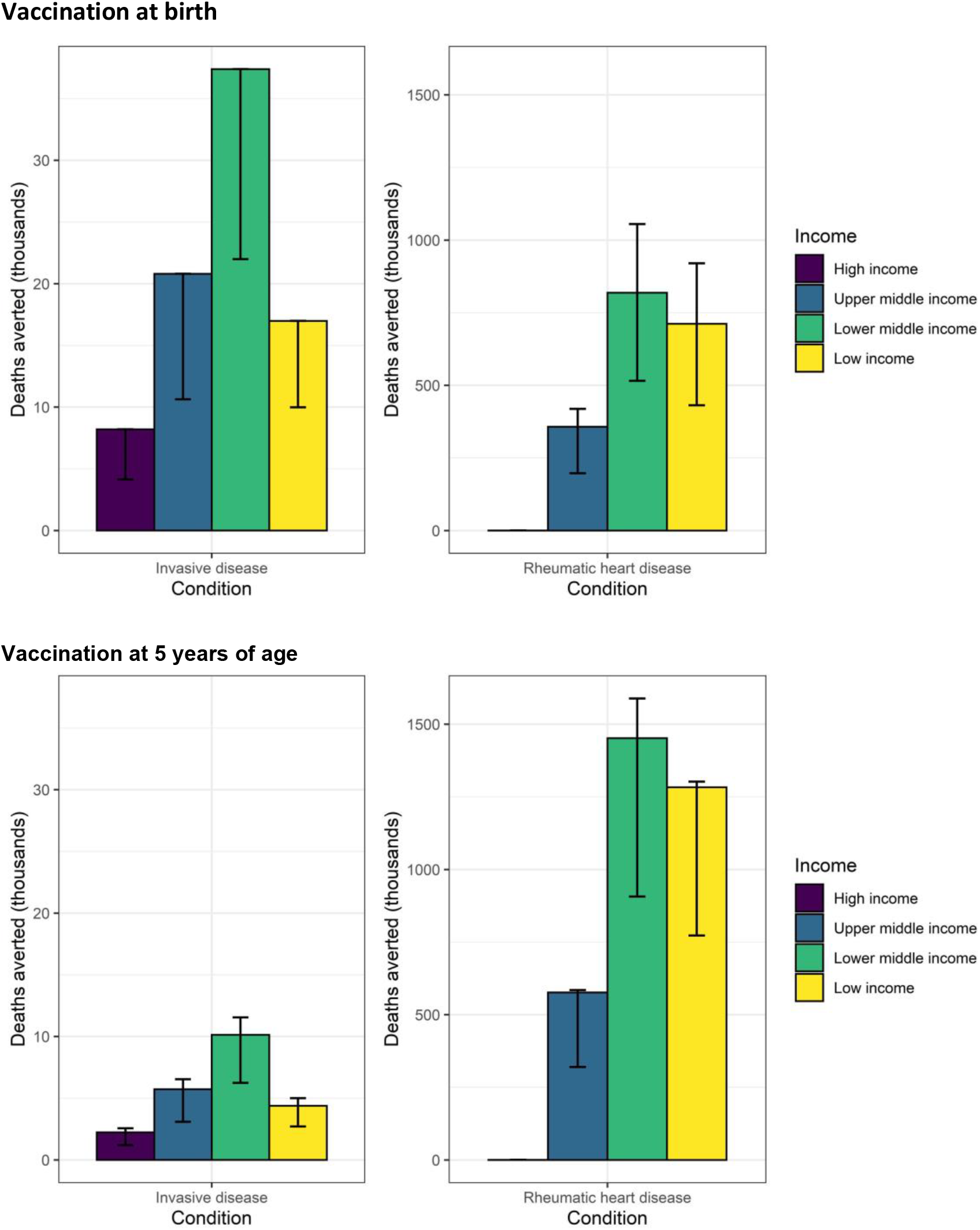
Vaccine impact (deaths averted) at the country-income level. The vaccine impact on deaths averted (in thousands) is stratified by income levels of countries (World Bank income classification), based on the lifetime health impact of vaccination at birth or 5 years of age for 30 vaccinated cohorts on Strep A disease burden (invasive disease and rheumatic heart disease). The vertical bars show the estimates for scenario 1, and the error bars show the range across scenarios 1-6. Note the differences in scale between the left panel (invasive disease) and the right panel (rheumatic heart disease). For rheumatic heart disease in high-income countries, the range of values for age 0 is (0.3, 0.6) and for age 5 (0.4, 0.7).

**Figure A3.**
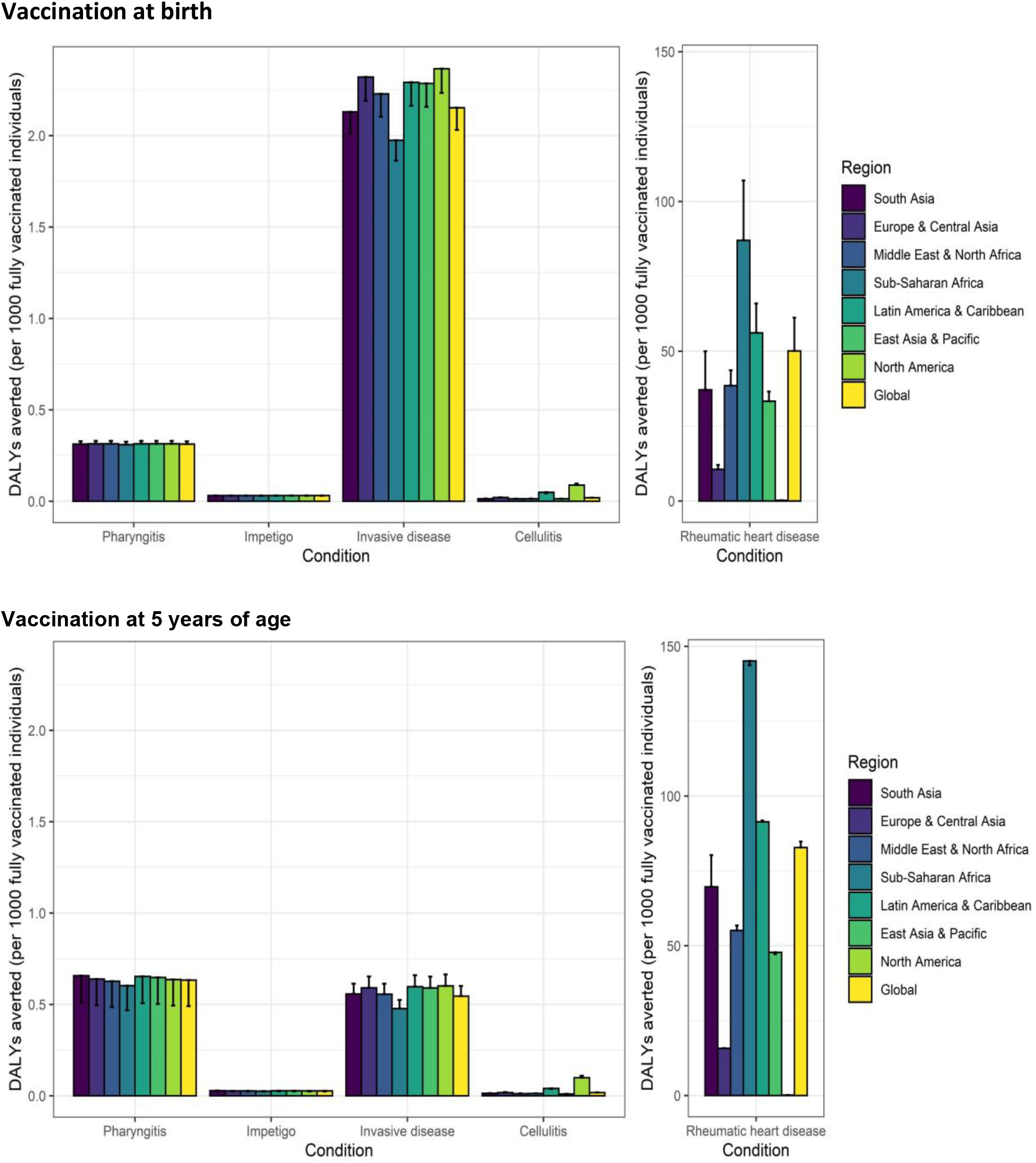
Vaccine impact (DALYs averted per 1,000 fully vaccinated individuals) at the regional and global levels. The vaccine impact on DALYs averted per 1,000 fully vaccinated individuals is stratified at the regional (United Nations regions) and global levels for different scenarios (estimate for scenario 1 and range across the six scenarios), based on the lifetime health impact of vaccination at birth or 5 years of age for 30 vaccinated cohorts on Strep A disease burden (pharyngitis, impetigo, invasive disease, cellulitis, and rheumatic heart disease). The vertical bars show the estimates for scenarios 1, 3, and 5 (which are equal), and the error bars show the estimates for scenarios 2, 4, and 6 (which are equal).

**Figure A4.**
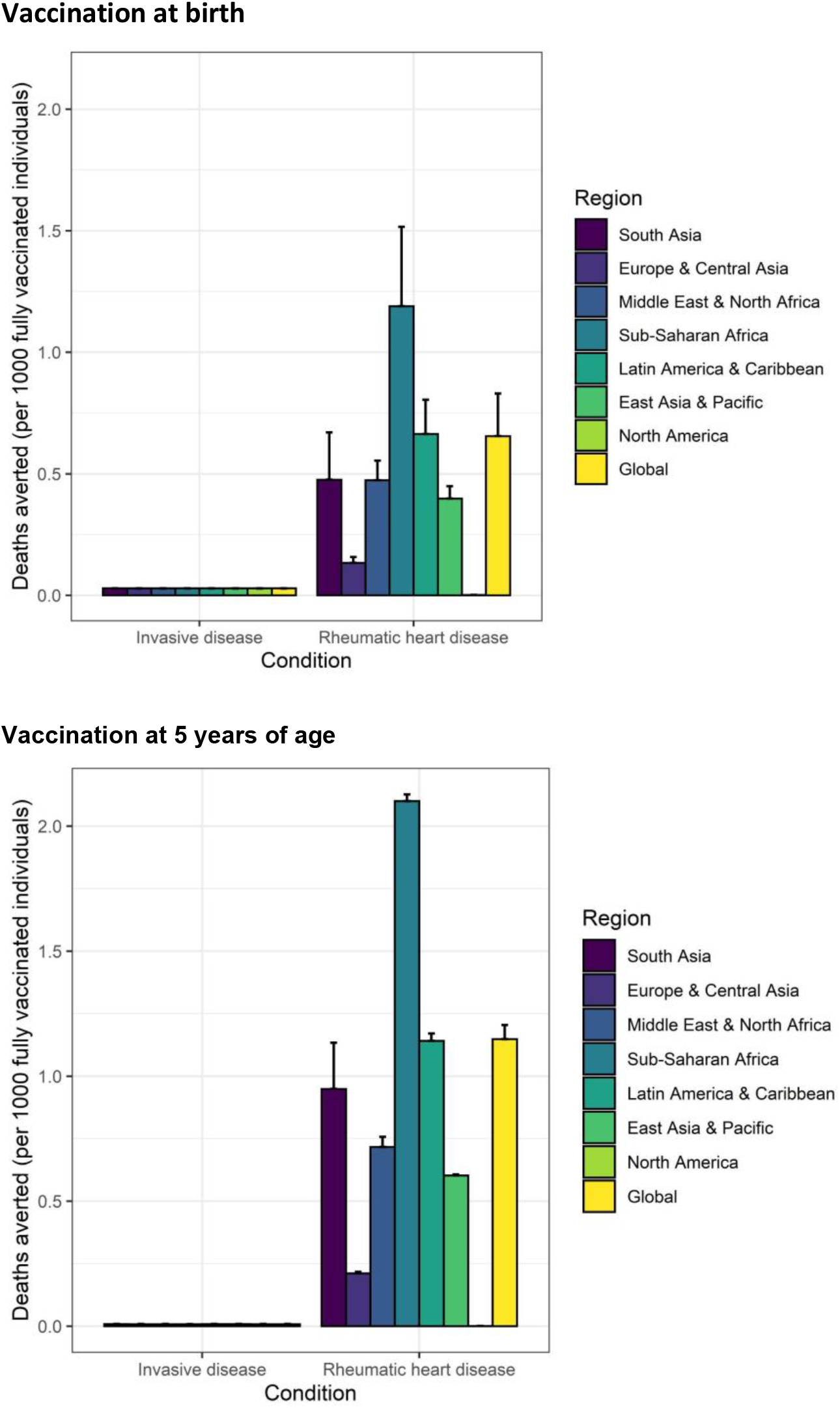
Vaccine impact (deaths averted per 1,000 fully vaccinated individuals) at the regional and global levels. The vaccine impact on deaths averted per 1,000 fully vaccinated individuals is stratified at the regional (United Nations regions) and global levels for different scenarios (estimate for scenario 1 and range across the six scenarios), based on the lifetime health impact of vaccination at birth or 5 years of age for 30 vaccinated cohorts on Strep A disease burden that results in death (invasive disease and rheumatic heart disease). The vertical bars show the estimates for scenarios 1, 3, and 5 (which are equal), and the error bars show the estimates for scenarios 2, 4, and 6 (which are equal).

**Figure A5.**
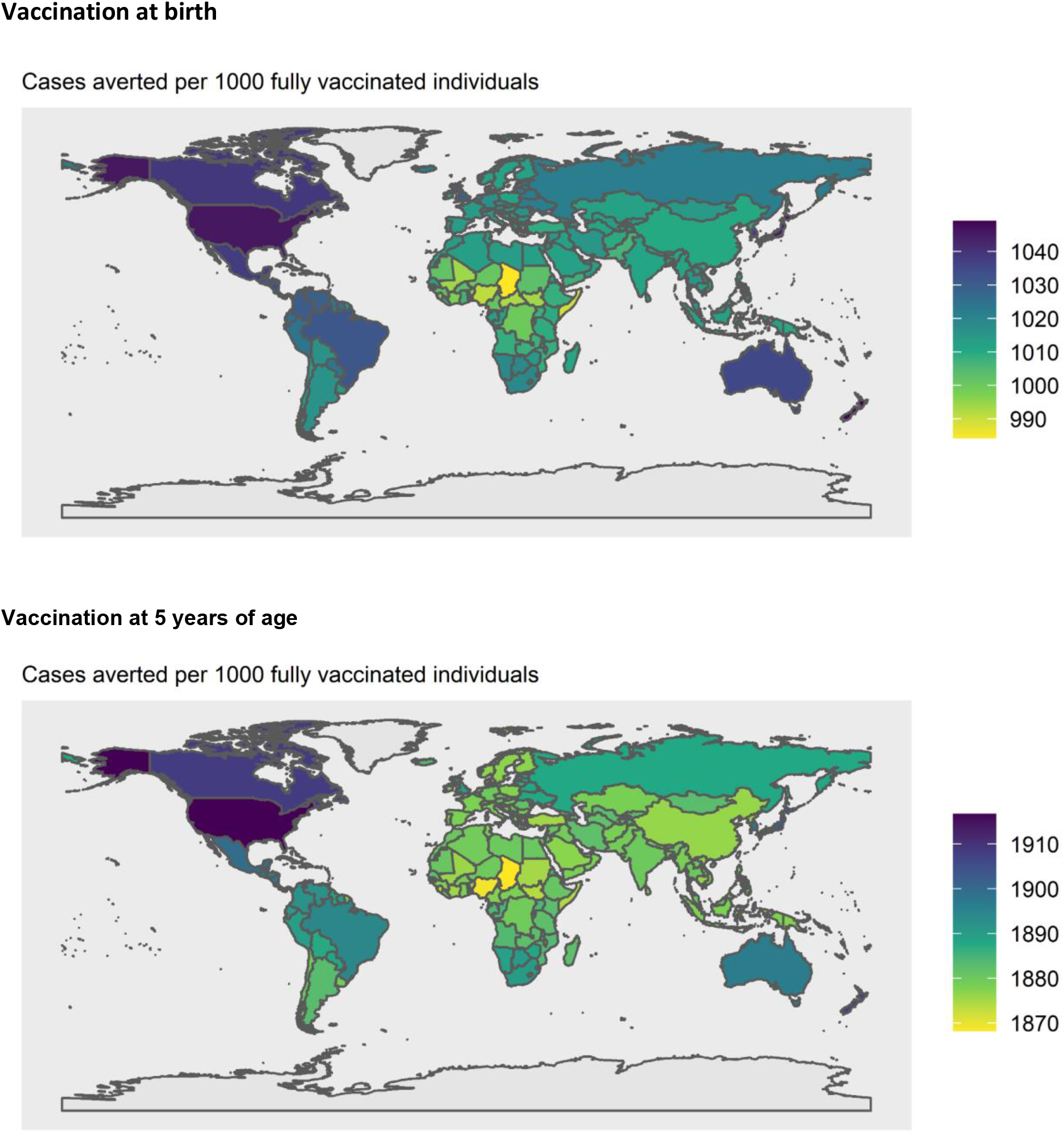
Vaccine impact (cases averted per 1,000 fully vaccinated individuals) at the national level. The vaccine impact on cases averted per 1,000 fully vaccinated individuals is shown for 183 countries, based on the lifetime health impact of vaccination at birth or 5 years of age for 30 vaccinated cohorts on Strep A disease burden (pharyngitis, impetigo, invasive disease, cellulitis, and rheumatic heart disease) for scenario 1.

**Figure A6.**
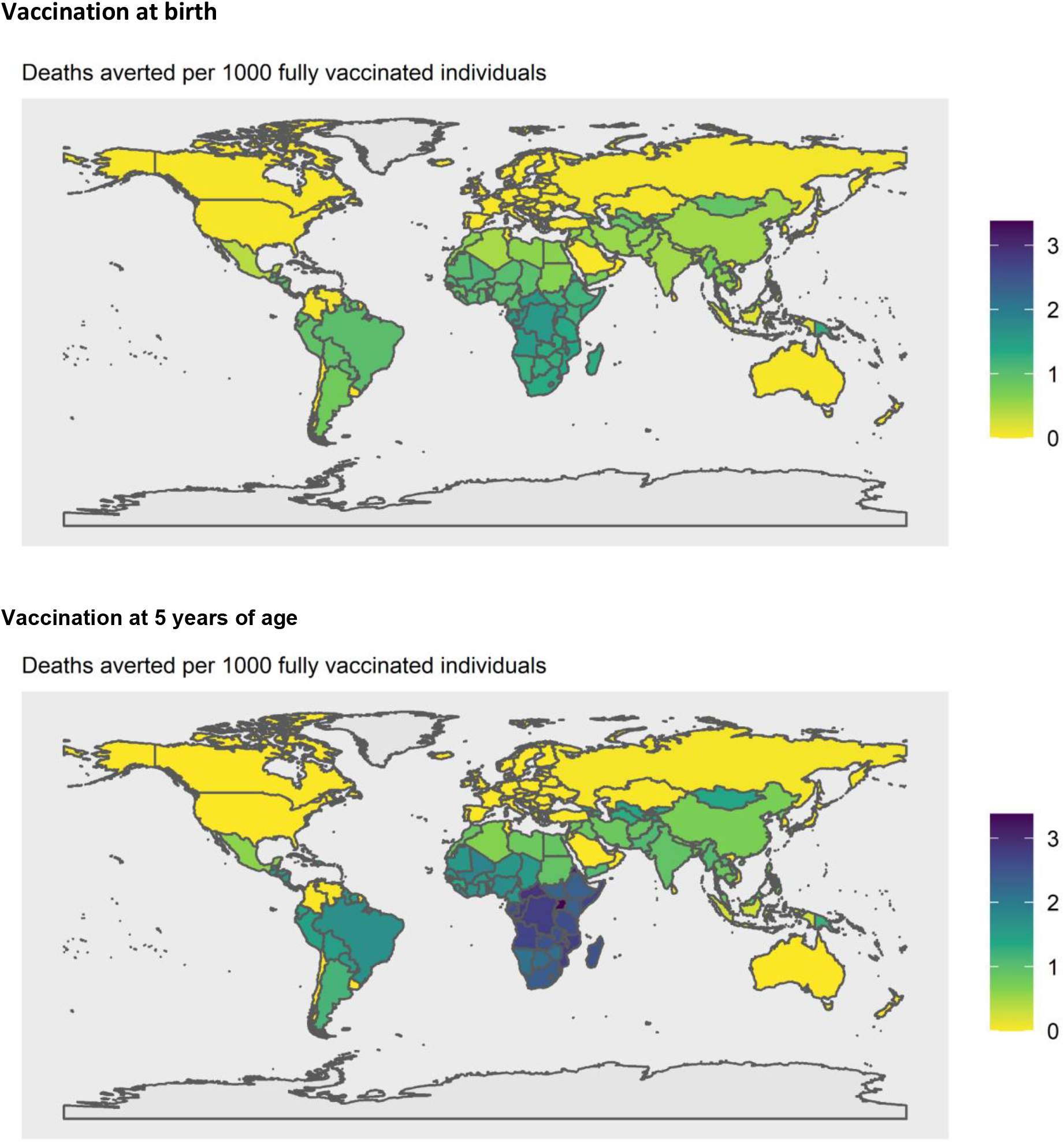
Vaccine impact (deaths averted per 1,000 fully vaccinated individuals) at the national level. The vaccine impact on deaths averted per 1,000 fully vaccinated individuals is shown for 183 countries, based on the lifetime health impact of vaccination at birth or 5 years of age for 30 vaccinated cohorts on Strep A disease burden resulting in death (invasive disease and rheumatic heart disease) for scenario 1.

